# An LLM-Based Comparison of Ambient AI Scribes for Clinical Documentation

**DOI:** 10.1101/2025.06.24.25330085

**Authors:** Jaison Jain, James Kaan, Suraj Jain, Austin Young, Camilo Martinez, William Kartsonis, Carlos Ortiz, Ryan Cheng, Erik Jaklitsch, Srinivas Cherukuri, Aleksandra Qilleri, Apostolos Tassiopoulos

## Abstract

Ambient AI scribes have become an increasingly promising option for automating clinical documentation, with dozens of enterprise solutions available. It remains uncertain whether models with domain-specific tuning outperform naïve models “out of the box.” This study evaluated five commercial AI scribes, alongside a custom solution using the base model of GPT-o1 without fine-tuning, as well as an experienced human scribe, in a series of simulated clinical encounters. Generated notes from these parties were scored by large language models (LLMs) using a rubric assessing completeness, organization, accuracy, complexity handling, conciseness, and adaptability. Our naive solution achieved scores comparable with industry-leading solutions across all rubric dimensions. These findings suggest that the added value of domain-specific training in ambient AI medical scribes may be limited when compared to base foundation models.

## Introduction

Documentation of clinical encounters is a time-consuming component of healthcare delivery contributing to physician burnout and decreased patient interaction (Shanafelt et al, 2017; Sinsky et al, 2016). Human scribes have long been employed to alleviate this burden on clinicians, with studies showing improvements in RVUs per hour, provider satisfaction, and patient satisfaction (Gottlieb et al, 2020). Despite their benefits, human scribes are expensive and limited by inconsistent quality, staffing shortages, and high turnover. Ambient AI scribes have emerged as promising tools to accelerate and potentially improve documentation while addressing the limitations of human scribes (Davenport & Kalakota, 2019). Numerous solutions are currently available in the market; some established almost a decade ago, while others emerged within the past year. In the current environment of rapidly improving AI capabilities, it is unclear whether long-standing solutions, boasting custom model refinements and fine-tuning on thousands of real-world patient encounters, outperform base versions of the latest AI models. The expanding breadth of knowledge in the most recently developed AI models, combined with the advent of “reasoning” capability (e.g., GPT-o1), underscores the possibility of the newest models being industry-competitive without refinement. To explore this question, we tested a series of commercial AI scribes alongside our naive solution (termed “Om”) on simulated cases designed to reflect the diversity and complexity of real-world clinical settings.

## Methodology

This study was conducted at the Stony Brook University Renaissance School of Medicine (RSOM) Simulation Center with volunteer participants enacting six clinical scenarios: a simple primary care case, complex primary care case, psychiatric encounter, post-operative follow-up, trauma case, and inpatient encounter. Each scenario involved scripted dialogues that were recorded simultaneously by devices running AI scribe applications. Five commercially available scribe systems (Scribes A-E) were tested, along with an experienced human scribe with over 5 years of scribing experience in a professional clinical setting (Scribe F), as well as our solution (Om). Case dialogue was designed to reflect the variation and unpredictability of real-world clinical encounters.

To evaluate the generated clinical notes, we developed a rubric assessing notes on six key dimensions: completeness and relevance of clinical content; organization and clarity; accuracy and specificity; handling of complexity and interruptions; conciseness and readability; and adaptability to varied clinical workflows. Rather than relying on human evaluators for scoring, we employed LLMs to score clinical notes—namely OpenAI o1-pro, Grok 3, Claude 3.5, and DeepSeek R1. These models represented the latest reasoning capabilities at the time of writing. LLM-based evaluation was chosen over human-based evaluation to reduce evaluator bias, provide reproducible assessments, and generate results time-efficiently. Follow-up studies with human surveys are being planned. Each LLM evaluator was prompted with the case dialogue, the evaluation rubric, and the corresponding scribe-generated notes from each participant. Participants were anonymized to all model evaluators. The model then returned a structured object representing scores across all rubric criteria. Data was aggregated and processed in R for statistical analysis and visualization.

## Results

Our untrained solution (Om) consistently achieved first-rank scores across OpenAI o1-pro, Claude 3.5, and Grok 3 (4.64, 4.69, and 4.72, respectively), and ranked second (4.75) behind Scribe A (4.86) under DeepSeek R1’s evaluation. Om was thus ranked first by three of the four LLM evaluators (Figure 1).

**Figure 1.**
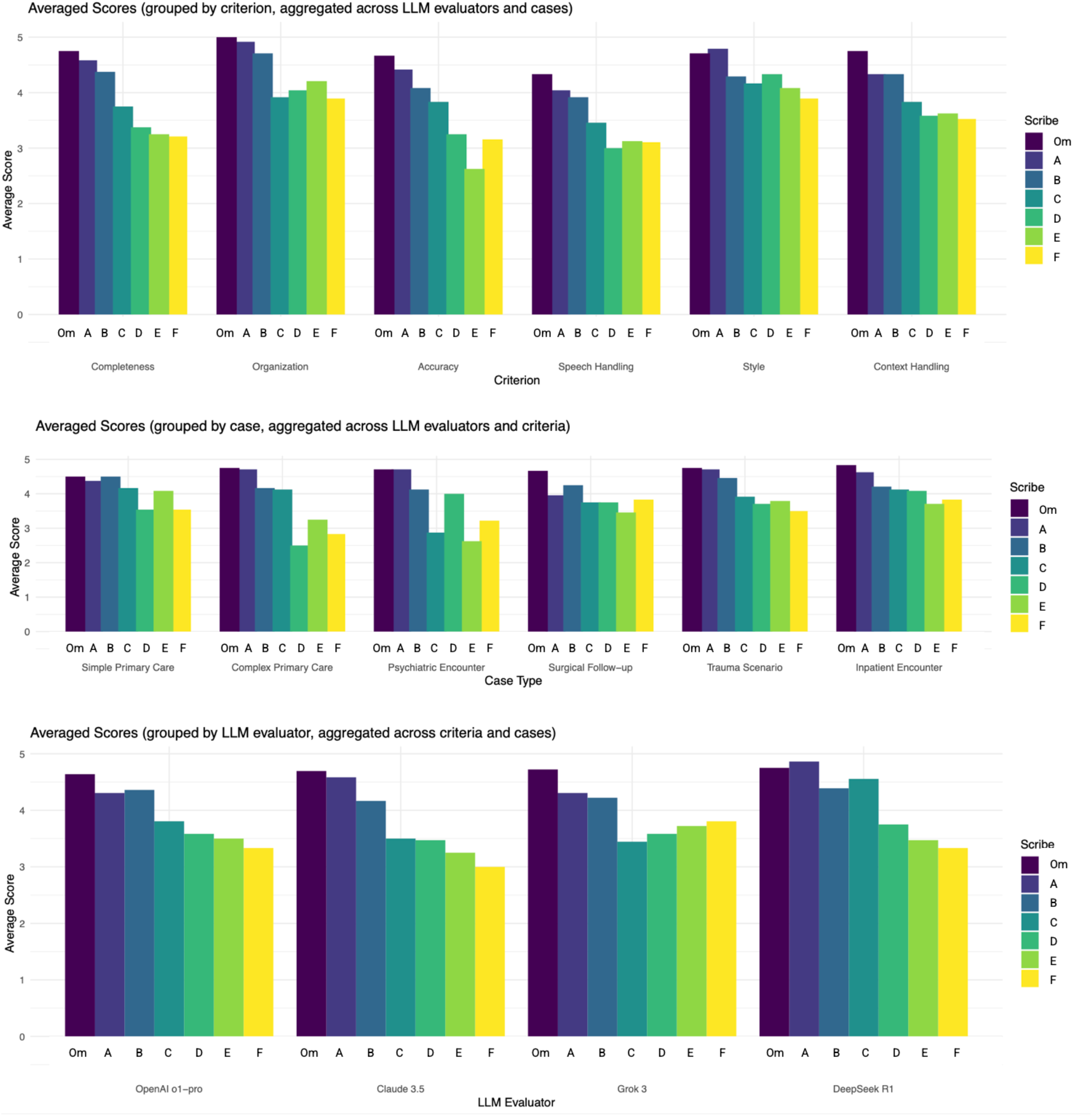
Scribe Performance by LLM Evaluator, Criterion, and Case. (a) Average scores grouped by large language model (LLM) evaluators. (b) Scores aggregated across rubric criteria. (c) Scores aggregated across clinical scenarios. Scribe F = human scribe.

Analysis by rubric criterion revealed that Om led in Completeness (4.75), Accuracy (4.67), Organization (5.00), Context Handling (4.75), and Speech Handling (4.33). Scribe A exceeded Om in Style (4.79 vs. 4.71). Om maintained the highest composite scores across all other metrics. Case-specific analyses show that Om held the highest or jointly highest score in all six scenarios, ranging from a mean of 4.50 in simple primary care to 4.83 in the inpatient encounter (Figure 2).

**Figure 2.**
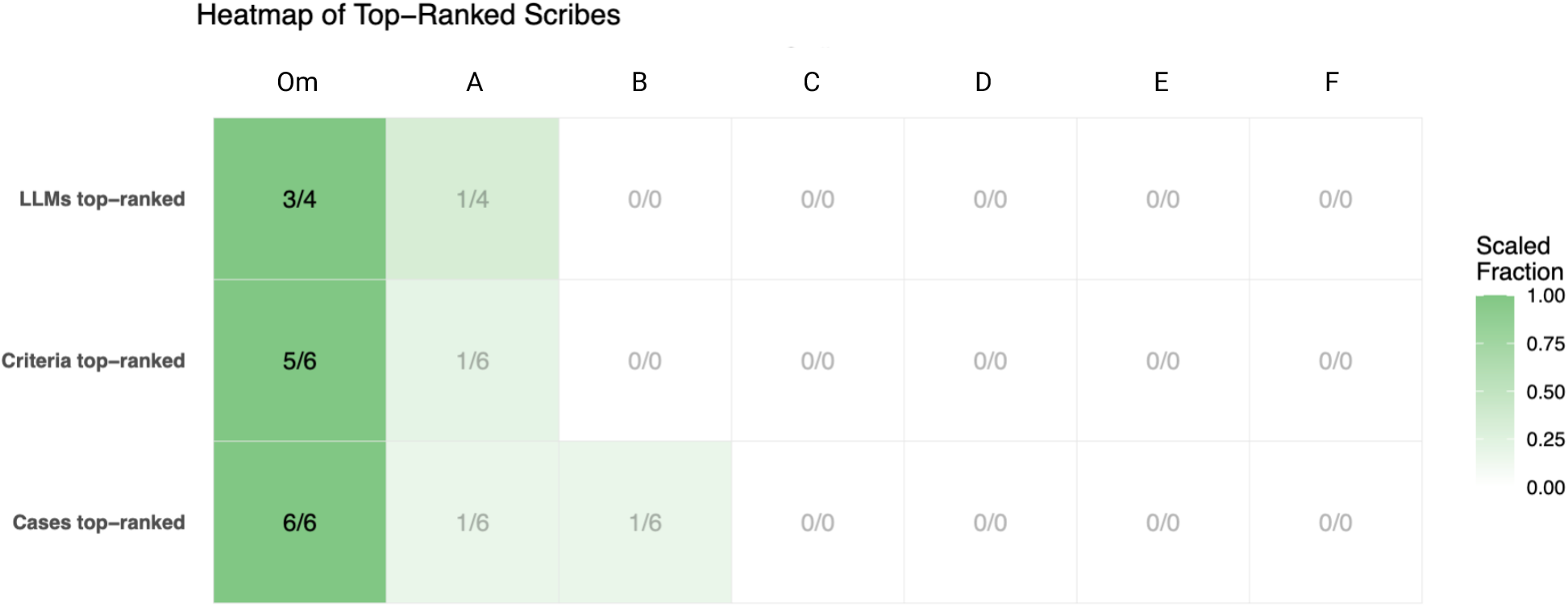
Frequency of Top-Ranking Across Scribe Evaluations. Heatmap summarizing how often each scribe achieved the top ranking across evaluations by LLM evaluator evaluation criterion, and clinical case. Darker shading indicates a higher proportion of top ranks. Scribe F = human scribe.

LLM evaluators with “reasoning” ability (DeepSeek R1 and OpenAI o1-pro) exposed their chain of thought and could therefore be introspected to understand the difference in scores. Notable details mentioned by the LLM were as follows: inaccurate transcription of medical terms (e.g, Scribe D replaced “IV” with “MIV” in its note for case 1), misstatement of patient age by Scribe E (e.g, reports patient’s age as 73 rather than 20 in case 3), and inability to deal with inconsistency (e.g, EMS reported left arm IV placement whereas nurse reported right arm placement in case 5; all scribes arbitrarily chose a laterality whereas Om refrained from doing so).

## Discussion

The present study evaluated seven scribes in a range of simulated clinical scenarios using LLM-based evaluation. Participants included five well-known enterprise solutions, one experienced human scribe, and one custom-built solution utilizing the latest AI reasoning model (GPT-o1). Our results suggest that a naive solution—one built on the base model of an AI reasoning model with no further refinement or tuning—may exceed or at least match the performance of industry-leading solutions when scored with LLM-based evaluation. Om achieved top or near-top aggregate scores for completeness, accuracy, organization, context handling, and speech handling. Cases involving complex or nuanced clinical interactions (e.g., trauma, psychiatric) tended to showcase the importance of robust speech handling, context retention, and correct capture of clinical details, dimensions in which Om performed well.

Om’s competitive performance may stem from the significant reasoning capabilities and expanded knowledge base of the latest AI models. While established solutions may have undergone domain-specific tuning, our results implicate that these adaptations may also limit generalizability and result in unforeseen errors (e.g, misstated patient age). It is plausible that the rapid advancement of foundation models may be outpacing the benefits of domain-specific optimization, challenging conventional wisdom that extensive fine-tuning maximizes performance for clinical AI applications.

Several important limitations should be noted. First, no real-world evaluation was included to evaluate actual workflow improvements or reduction in documentation time. All notes were derived from a controlled setting using simulated cases. Second, we employed LLM-based evaluators rather than human evaluators to reduce the inter-rater bias inherent to human scoring. The broad agreement among multiple LLM evaluators in scribe rankings indeed supports the reliability of this methodology. However, LLM-based evaluation may not reflect the preferences or nuanced clinical judgement of a human evaluator. Future studies should, therefore, incorporate real clinical workflows, external physician panels, and robust time and cost analyses to assess the efficacy of these AI scribes more comprehensively.

## Data Availability

All data produced in the present study are available upon reasonable request to the authors

## Acknowledgements

This study was conducted at the clinical simulation center of Stony Brook University Hospital, Renaissance School of Medicine, with the help of volunteers including: Matthew Tharakan, MD, Lyncean Ung, MD, and Rachel Wong, MD.

## Data and Code availability

R code, case dialogues, generated notes, and raw score data are available upon request.

