## Supplemental Info for "An LLM-Based Comparison of Ambient AI Scribes for Clinical Documentation"

### Supplementary Information

#### 1. Rubric

The following rubric was used to score all AI-generated clinical notes:

1. Completeness & Relevance of Clinical Content
   a. History of Present Illness (HPI)
   Criteria: Does the note accurately capture the patient’s stated symptoms, their timeline, and any important contradictions or inconsistencies?
   Things to look for:
   Multiple symptoms mentioned, possibly “out of order.”
   Contradictory statements by the patient, or changes in the story.
   Additional complaints added late in the conversation.
   High-Quality Note: Thoroughly documents all symptoms, including any last-minute complaints or contradictory statements, in a coherent, chronologically logical manner.

b. Past Medical, Surgical, Social, and Family History
Criteria: Does the scribe capture relevant history that the patient mentions, including someone else’s health problems (if relevant to the plan)?
Things to look for:
Inclusion of any discussion about family members/caregivers’ health if relevant.
Social determinants of health (SDOH) factors (e.g., living situation, access to care, financial barriers) when they are mentioned.
High-Quality Note: Accurately includes all relevant historical elements, especially SDOH, in appropriate sections.

c. Review of Systems (ROS) & Physical Exam Findings
Criteria: Are the pertinent positives and negatives from the conversation captured?
Things to look for:
Clear delineation of symptoms associated with each body system.
Any exam findings (if performed or discussed) accurately transcribed.
High-Quality Note: Summarizes relevant positives and negatives without omitting or misrepresenting symptoms.

d. Medical Decision Making (MDM) & Assessment/Plan (A/P)
Criteria: Are the clinician’s thoughts, reasoning, and intended next steps accurately documented?
Things to look for:
Specific mention of tests, referrals, or treatments discussed.
Rationale behind each medical decision included (when expressed by provider).
Presence of an organized Assessment/Plan section that ties symptoms and findings together.

High-Quality Note: Clearly outlines the reasoning (MDM) and well-structured A/P that reflects all problems discussed (including late additions).

2. Organization & Clarity
   a. Logical Flow
   Criteria: Do the notes follow a recognizable clinical documentation structure (e.g., CC/HPI, ROS, Exam, MDM, A/P) despite the conversation being "out of order"?
   Things to look for:
   Whether out-of-sequence patient statements are coherently reorganized.
   High-Quality Note: Presents information in a standard clinical order or at least a clearly labeled structure that is easy to follow.

b. Separation of Non-Medical Verbiage
Criteria: Does the note exclude irrelevant small talk (weather, sports, etc.) while retaining relevant psychosocial factors or SDOH?
High-Quality Note: Filters out unrelated content but keeps any personal or social factors pertinent to the care plan.

3. Accuracy & Specificity
   a. Proper Terminology & Correct Detail
   Criteria: Are medical terms used correctly? Are lab values, medication names/doses, and patient instructions accurate?
   High-Quality Note: Uses appropriate clinical language; no errors in dosage, medication names, or major details.

b. Managing Contradictory Statements
Criteria: How does the note handle patient contradictions? Does it document them objectively without creating confusion?
High-Quality Note: Objectively records contradictions with clarifying statements or notes uncertainty where applicable.

4. Handling Complexity & Interruptions
   a. Multiple Speakers / Interjections
   Criteria: Are statements from caregivers, parents, or other interrupters captured accurately and attributed appropriately?
   Things to look for:
   Clear attribution of who said what (e.g., patient vs. parent).
   Correct capturing of context for interruptions or additional commentary.
   High-Quality Note: Maintains clarity about which speaker is providing information.

b. Dealing with Rude or Unprofessional Tones
Criteria: Does the note objectively capture the situation without inserting personal bias or unnecessary editorializing?
High-Quality Note: States facts neutrally (e.g., "Patient used harsh language regarding wait time") without inflammatory or judgmental wording.

5. Conciseness & Readability

a. Length & Brevity

Criteria: Is the note concise yet complete?

High-Quality Note: Avoids excessive repetition while including all pertinent details.

b. Grammar, Spelling, and Formatting

Criteria: Is the documentation free from grammatical, typographical, and formatting errors?

High-Quality Note: Reads smoothly with minimal errors, uses headings or bullet points effectively where needed.

6. Adaptability to Varied Clinical Workflows

a. ED, Inpatient, and "Visit Hopping" Scenarios

Criteria: Does the AI adapt the documentation format for different settings (e.g., ED admission vs. inpatient discharge summary) or multiple simultaneous patient discussions?

High-Quality Note: Presents coherent notes tailored to the clinical setting; if "visit hopping" occurs, the correct details end up under the correct patient/encounter.

b. Pre-Charting & Post-Charting Annotations

Criteria: Are any pre-visit or post-visit additions (like relevant labs or discharge instructions) handled and labeled correctly?

High-Quality Note: Makes it clear which parts of the note are from prior documentation vs. the current encounter or the follow-up.

7. Scoring / Rating Scales (Example)

RUBRIC:

Completeness & Relevance

1: Many missing elements, important complaints not captured

3: Most key elements present, minor omissions

5: Thorough and accurate, includes all relevant details

Organization & Clarity

1: Disorganized, no recognizable structure

3: Some logical structure, mild confusion in ordering

5: Well-structured, easy to follow

Accuracy & Specificity

1: Multiple clinical inaccuracies or incorrect terminology

3: Mostly correct with minor errors

5: Highly accurate and uses appropriate clinical language

Handling Complexity & Interruptions

1: Misses or confuses different speakers; does not handle abrupt changes

3: Captures interruptions and multiple speakers with some clarity issues

5: Clearly differentiates speakers and conversation threads

```
Conciseness & Readability
1: Verbose, repetitive, or unclear grammar
3: Generally readable, some repetition or minor errors
5: Concise, clear, with minimal errors

Adaptability to Workflows
1: Fails to adapt to ED vs. inpatient context, merges patient
data incorrectly
3: Works for one context well but struggles with others
5: Shows robust adaptability and clear labeling for each scenario
```

##### 2. Scoring prompt

The following prompt was used by LLM evaluators in scoring clinical notes (the "{rubric}" placeholder refers to the text of the rubric above, "{transcription}" is speaker-labelled transcription of the simulated medical encounter, and the "{notes}" placeholder refers to a concatenation of all notes to be evaluated).

```
Below is a transcription of a medical encounter, followed
by a series of clinical encounter notes generated by
anonymized AI or human scribes.

{transcription}

{notes}

Please evaluate these notes according to the following
rubric:
{rubric}

Please output the evaluations of each note in the following
format:
{<company name>: {<criteria index 1>:<criteria score 1>,
<criteria index 2>:<criteria score 2>, ...} , ...}
Score per criteria (eg '1') not subcriteria (eg '1b')
```

##### 3. Case dialogues

##### 3.1. Case 1

DOCTOR: "This is a 77-year-old male with a history of hypertension and diabetes visiting my office with the primary complaint of low back pain. I reviewed his lab work that was obtained recently and his hemoglobin A1c has increased from 6.9 to 8.6%."

DOCTOR: "Hi, Mr Bruno. It's nice to see you again. How's the family? Did your daughter have the baby? What brings you in today?"

PATIENT: “Yes…she is beautiful…5 pounds 6 ounces…She is such a good baby. She is not fussy, just living her best life, and she does not cry. The labor was only an hour long, and the hospital was great. My family is good. My wife Crystal and I just got back from vacation. It was great to be empty nesters. But my back is killing me.”

DOCTOR: “OK, when did it start?”

PATIENT: “Well, it’s complicated. I’ve always had occasional back pain, but this is something else it started during my trip to Spain.”

DOCTOR: “OK, were you doing anything when it started?”

PATIENT: “Yeah. So, during my trip last week, Crystal tried to make me eat healthy. I went to the local grocery stores, but I had to carry all these groceries and the hotel I was staying at was pretty far, so by the time I got to the hotel, I started noticing that I had some back pain.”

DOCTOR: “Did you take anything?”

PATIENT: “Not at first. I’m a pretty strong guy and can take a lot. I used to be the quarterback in high school. I just let it ride until the next day, and then Crystal forced me to take a Tylenol since I didn’t want to go to morning Yoga. Although I didn’t want to go, I took some.”

DOCTOR: “OK. Yeah, I can understand why you don’t like Yoga. I’m not too fond of it myself. Let’s return to your back pain; can you tell me more about it? Can you tell me where the back pain is and what it feels like?”

PATIENT: “Sort of around my entire lower back kind of really bad when I try to, you know, move around in any direction.”

DOCTOR: “OK. Do things like lying down or anything besides the Tylenol make it better?”

PATIENT: “Yeah, I guess now that you mentioned it when I lay down, it feels much better. The Tylenol did help a bit, but it still worries me.”

DOCTOR: “Any trauma or injuries during the trip?”

PATIENT: “Well, after the shopping thing, we were at the running of the bulls. I bumped into a bench and then I fell backward and hit my lower back.”

DOCTOR: “Oh no. Were you okay? Did you have any cuts or bruises? Did you get evaluated by anyone after that incident?”

PATIENT: “Of course I was okay. Did I mention that I was ALL COUNTY MVP in 1970? I only had a small bruise, but I used to get hit playing football all the time back in the day.” --

{Interrupt}
NURSE: “Excuse me, doctor. I know you don’t like interruptions, but you have a phone call from your mother, who said it is important.”

{Pause Ambient AI technology}

DOCTOR: “Please excuse me for a few minutes.”

{Doctor returns}

DOCTOR: “I am sorry about that. So, a lot has happened to you. I will need to ask a few more questions to understand your back pain a little more. How bad was your back pain when it started? Like 10 being the worst pain you've ever had, one being no pain.”

PATIENT: “I would say it's a 6. But I have a high pain tolerance, remember I was MVP ALL COUNTY 1970.”

DOCTOR: “OK. And after the Tylenol and resting it, how is the pain now?”

PATIENT: “It's probably like a four.”

DOCTOR: “OK, umm and did the pain ever go down the legs or kind of, you know, move anywhere else besides your lower back? And any changes in your bowel or bladder habits? Are you pooping the same are you peeing ok?”

PATIENT: “Umm, no, no. But since you mentioned it, the pain stayed in my back, but my little guy didn’t work as well as I hoped for during the trip. I am not sure if it is related, but I want to mention it.”

DOCTOR: “Do you mean that you were not able to have an erection?”

PATIENT: “Yes, I am a little embarrassed about it.”

DOCTOR: “How long have you had the erectile issues? Did you start having this issue after your back injury?”

PATIENT: “No, it happened a few years ago, but I have been embarrassed to mention it. I’d say it has been five years that I have not relations with my wife. Come to think of it, it’s more like 10 years. It has been affecting my self-esteem and possibly my marriage. Sometimes, I think Krystal might divorce me.”

DOCTOR: “I didn’t know that you were feeling this way. I want to talk more about this since it has such a negative impact on you. I also want to address the primary reason why you are here which is your back pain. Since your erectile dysfunction does not seem related to the back pain, do you mind if we hold this topic until the end of the visit?

PATIENT: “Sure.”

DOCTOR: “OK. Just remind me, how long ago did the back pain start?”

PATIENT: “Well, always had some chronic occasional back pain like a couple times per week. All great quarterbacks have it. Many quarterbacks have the same thing throughout their careers. This back pain is new and different and started two weeks ago in Pamplona.”

DOCTOR: “Was it after plane ride back from Spain?”

PATIENT: “Nah, it started with the groceries, the economy seats in the plane didn’t help none. I am sure it is more comfortable in first class.”

DOCTOR: “Actually, I fly economy. More importantly, aside from Tylenol, have you taken any other medicines like Advil, Aleve, naproxen, that kind of thing?”

PATIENT: “No. They tried the other medications, and since it didn’t work, my cousin Bobby gave me one to two of his Percocets. That worked!”

DOCTOR: “I understand that you were in a lot of pain at the time, but it is important not to take other people’s prescribed medications. Percocets can be addictive and dangerous because they cause depression with breathing. And have you had any like back surgeries, any back problems in the past?”

PATIENT: “No back surgeries, just the occasional back pain every now and then. That's why this worries me this pain is different.”

DOCTOR: “Any unintentional weight loss?”

PATIENT: “Since the vacation, I have gained some weight. I had too much sangria and tapas”

DOCTOR: “OK, alright, alright. I'm going to examine you for a second. I also was just gonna follow up on a couple of things. Umm, it's seems that, you know, with the diabetes you were doing really well last time.
Has that been OK? Especially with your travels to Spain and eating healthy, have the sugars been alright?”

PATIENT: “I mean the paella was irresistible and it paired well with the sangria, so I couldn't help myself. So I guess I cheated a little bit on my diet, that’s what happened. Crystal was on my case about that, and the money we were spending, so then I had to go grocery shopping, and now here I am.”

DOCTOR: “We all cheat with our diets but have you been checking your sugars?”

PATIENT: “No, it's just I wanted to pack light because I knew it was gonna walk around a lot. So, I didn't bring my glucometer with me.”

DOCTOR: “OK, alright, cause I actually see on the latest set of labs, your hemoglobin A1c was a bit higher like 8.6%. When were those labs done? I think it was before your trip, lemme look, oh that was done yesterday. So maybe we could talk about that in a little bit too. Alright. And anything else been going on?”

PATIENT: “Well, my penis doesn’t work, I injured my back, and now you are telling me my diabetes is uncontrolled. I think that’s enough.”

DOCTOR: “I know it’s a lot, I understand, but we will get through this together, you’ll be fine. Let's go ahead and do an exam. I'm gonna take a look at your back to do a full exam. OK, it looks like your blood pressure is pretty good 118 / 70. And it seems like your weight is stable at 156 pounds. I’m going to speak out my physical exam findings so my app can help me write this note. You are awake alert and oriented, Tongue is midline. Pupils equal and reactive. Heart regular rate and rhythm, no murmurs rubs or gallops. Lung sounds are clear to auscultation bilaterally. Does it hurt when I push on your belly?”

PATIENT: “No, no.”

DOCTOR: “Abdomen soft and non-tender in all four quadrants. Bowel sounds present times 4. Now I’m going to examine your back. Any pain right now?”

PATIENT: “No, it feels fine.”

DOCTOR: “No. OK, nice. I am feeling right here on your back on your spine right now. Are you feeling any tenderness when I press here on your lower spine?”

PATIENT: “No, no, nothing, nothing there.”

DOCTOR: “OK. And now I'm going to press on either side. There are those muscles called your paraspinal muscles. I’m pressing on them. Do you feel any pain?”

PATIENT: “Owww, now that hurts. Stop…”

DOCTOR: “Oh, OK, alright. I'm not going to keep touching, but like, is that the main area where you're having pain?”

PATIENT: “Yeah, that's it.”

DOCTOR: “OK. And to be thorough I’m going to just test the strength in your legs as well. OK. Can you push with both legs over here for me? And over here.”

PATIENT: “Yes.”

DOCTOR: “Alright, your reflexes and your strength in both lower extremities seem good and you're not losing any sensation in both legs. So that's a good sign. Umm, let's talk about a plan. It seems like you have a muscle strain in your back. Those muscles that I pressed on, that paraspinal muscle, that one seems really tender. And especially with it hurting when you move around, I think you probably just strained that muscle. In terms of the Tylenol that is a good medicine for you because it won't raise your blood pressure. It won't mess with your kidneys or give you any stomach issues, so that one you could take up to a maximum of two tabs of 500 milligrams, three times a day. I'm going to write that down for you, OK?”

PATIENT: “Can you give me a few Percocets since it helped me recently?”

DOCTOR: “Well, I think you haven’t given over-the-counter medication a try yet. You can go up to 1000 mg of Tylenol 3 times day, and some Salonpas Patches which have lidocaine in them, you can get them at Walgreens.” That should control the pain, it'll slowly get better. You can also put a little bit of like heat on the back and that will help and I’d imagine over the next week or so it's gonna be much better.  The 1970 QB MVP can get over this without resorting to opiates. If it doesn’t work, then we can revisit the topic, maybe do some imaging. I'd avoid resting in bed like fully, but you, you know, so just do light activities. Take it easy. I wouldn't do your normal gym exercises right now. And also if it's still there, we can consider a round of anti-inflammatories like an ibuprofen, naproxen kind of thing.”

PATIENT: “I’ve been reading online about acupuncture. Can I try it? There is an old lady down the street from me who does it, and I read online that it works.”

DOCTOR: “Sure. That's absolutely fine. Umm, I don't think there's any harm in it. And yeah, you could go ahead and try that as well. Alright. And then also you know if that Tylenol isn't working, you can you could go ahead and take like two of the naproxen over the counter twice a day with food. So those are sort of like your backup plan. If the Tylenol and the heat and the rest aren't taking care of it, we could touch base. Let me know on the patient portal if you're still having symptoms. I did want to spend just a minute talking about your diabetes. Based on your latest hemoglobin A1C, your sugars are a little high. It was 8.6% and the number we're looking for is less than 7% for HBA1c. So that means I probably have to talk about lifestyle and medication change. You're on the metformin. Have you been OK with that?”

PATIENT: “Yeah. I have been taking it daily, so I am surprised my numbers look worse.”

DOCTOR: “So, your body likes to have a balance with everything, including your sugar levels. When you eat large amounts of sugar-containing foods sporadically, it can cause spikes that can be represented on a blood test called the hemoglobin A1c. It measures your sugar levels over a period of time rather than at a spot check. When your sugars are not balanced long-term, it can cause other problems with your body. For example, it can affect your kidneys, heart, and nerves. I suspect that it is the cause of your erectile dysfunction. Does that make sense?”

PATIENT: “Yeah. Man, I hope it is not too late to fix this.”

DOCTOR: “Let’s start fixing it then. I think what we could do is raise it up to 1000 milligrams twice a day, and we can refer you to a diabetic educator to help with understand the disease and how you can monitor your sugar levels and stay compliant. Since you have 500’s now at home you could take two tabs twice a day until your meds finish, and then for the next prescription I'll send 1000 milligrams tabs. So it'll be twice the strength. So if you want to finish out those tabs at home, I'll send a new one and then I want you to recheck your hemoglobin A1c in three months. OK, from starting the new dose, I'm also gonna put in a prescription for like a urine and some of the other yearly things that we do for your diabetes. OK, everything else is pretty much up to date with your foot and eye exam, so I'm going to put those labs in for three months from now."

PATIENT: "OK. I get it."

DOCTOR: "Returning to your intimacy issues and relationship with your wife. Do you feel like she is going to divorce you? Are you feeling depressed?

PATIENT: "No, I don't think she is going to divorce me, and I am not depressed. I want to fix this problem, though."

DOCTOR: "Do you still get morning erections?"

PATIENT: "No doc, it's been a while…"

DOCTOR: "OK, no problems. I'll send labs, including a morning testosterone level, TSH, and Lipid panel, along with your A1c, and we can review the results at our next visit. Remember, uncontrolled diabetes can worsen nerve damage and contribute to this problem, so cut down on alcohol and listen to Crystal, and eat well. If the labs come back abnormal, we can refer you to urology."

PATIENT: "OK. Should I get those done before I come and see you again?"

DOCTOR: "Yeah, let's schedule a follow-up visit for about a week after the labs, OK?"

PATIENT: "OK. Thank you so much."

DOCTOR: "All right. You're welcome. I'll see you in three months. OK. Bye."

#### 3.2. Case 2

[Case text is long and thus not reproduced here. Please email corresponding author and case text will be made available.]

#### 3.3. Case 3

DOCTOR: "Hello, Mr. Jones, I'm Dr. Patel. How are you doing? How was your dinner?"

PATIENT: "It was cold."

DOCTOR: "I'm sorry to hear that. I will ask them to reheat it next time. I would like to talk with you to try to figure out what is going on and see how we might be able to help you. Can you tell me what brought you into the hospital today?"

PATIENT: {Talking rapidly and gets up from chair to pace} "I'm fine! You don't understand, I'm on a big mission! The Deep State, they need me to fix everything, otherwise everyone will be in trouble!"

DOCTOR: "It sounds like you have a lot on your mind. Can you tell me a bit about what's been going on recently? Why were the police involved?"

PATIENT: "They don't know the real threat! I was just trying to protect everyone! They don't understand the bigger picture! I have so much to think about. I’m trying to save the world here! It is too hot here; don’t you have air conditioning?”

DOCTOR: “It sounds like you have a lot of thoughts going through your head and that can be difficult at times. Have these thoughts been affecting your mood recently?”

PATIENT: “My mood, it’s fantastic! I’m the happiest I’ve ever been. But then, no one understands, no one gets it. Why don’t they see?”

DOCTOR: “Who are you referring to?”

PATIENT: “Them, they are the ones that keep me up at night?”

DOCTOR: “Are you having any difficulty sleeping?”

PATIENT: “Oh, I don’t need sleep! I’ve got so much energy. I spend my nights working on developing a video game and painting my house while I try to accomplish my mission during the day.”

DOCTOR: “I see that you have a lot of things going on. Do you ever feel overwhelmed?”

PATIENT: “Not at all, I sleep all day.”

DOCTOR: “You said you were developing video games all day and accomplishing your mission; how do you manage to sleep?”

PATIENT: {Shouting} “Why would I sleep all day? I’m invincible! I’m going to save the world. But I’ve been really angry, though. Those people at home don’t listen. They make me so mad. They don’t understand.”

DOCTOR: “Are you referring to your parents?”

PATIENT: “I don’t know who they are.”

DOCTOR: “Have you’ve felt like acting on those feelings of anger?”

PATIENT: “They push me too far. But I don’t want to hurt anyone. I just want them to see, they need to listen. I am doing this for them.”

DOCTOR: “Have you had any thoughts of harming yourself or others?”

PATIENT: “Of course, not.”

DOCTOR: “Understood. Let’s discuss your past psychiatric history. Have you been admitted to a psychiatric hospital before?”

PATIENT: “I’ve been in and out of hospitals. I was just in one 2 weeks ago. They just don’t get me. I’ve had highs, I’ve had lows, but no one understands what’s really going on.”

DOCTOR: “Can you tell me about any previous medications you’ve taken and if there were any issues with them?”

PATIENT: {Starts walking out of the room} “Medications! I don’t need medications, they don’t help! They make me horrible and dumb. I’m done talking.”

DOCTOR: “Mr. Jones, please come back to the room. It is important that we finish our conversation.”

PATIENT: “Whatever, you got to make this quick. I have more important things to think about.”

DOCTOR: “I’ll try to finish this up as quickly as I can. Do you have any medical issues or have you had any surgeries?”

PATIENT: “No, surgeries. Just the mission! My health is fine! But they keep sending me to these hospitals. They don’t understand.”

DOCTOR: “Do you have any history of violence?”

PATIENT: “I’m not violent. I’m protecting people. (Whispering) Did you see that light? I think they are trying to communicate with me?”

DOCTOR: “I didn’t see any light. Who is trying to communicate with you?”

PATIENT: “I’m done. I need to go to pee.” {Patient leaves the room}

{Post-charting}

DOCTOR: “Patient is 20 y/o male with history of bipolar disorder and no significant medical history with multiple past inpatient psychiatric hospitalizations (last bring 2 weeks ago), no established outpatient treatment, non-compliant with medications and no know past suicidal behavior who was brought in by police department from home secondary to erratic behavior. Patient presents with symptoms of mania including labile mood with increased goal directed activity, racing thoughts, decreased need for sleep and grandiose/paranoid delusions. Patient denies any auditory or visual hallucinations. He does not report any symptoms of depression at this time. He denies any suicidal or homicidal ideation, intent or plan. Patient at this time not able to care for self and would benefit from inpatient psychiatric hospitalization for further stabilization.

Although it is less likely to be these other differentials, I must consider Drug intoxication (Cocaine, Methamphetamine, Alcohol), Overdose (Acetaminophen, Salicylate), Thyroiditis, Thyrotoxicosis, and Electrolyte derangements (hyponatremia). I will need to obtain a urine drug screen for possible illicit drug use. I will also need an EKG, CBC, Chemistry, Liver Function Panel, Acetaminophen Level, Salicylate Level, Ethanol Level, and Thyroid Function Panel.

I will initiate the patient on medication for management of manic/psychotic symptoms while expanding collateral and encouraging patient to engage in therapeutic milieu.”

##### 3.4. Case 4

DOCTOR: “I will be seeing this 45-year-old male who I performed a cholecystectomy on one week ago. He was prescribed oxycodone for his pain. He has no significant medical history or allergies to medications. He did not have any other surgeries.”

{Enter into patient’s room}

DOCTOR: “Good afternoon, Mr. Stone. How are you feeling?”

PATIENT: “I am feeling okay and I would like to know if I can go back to work soon.”

DOCTOR: “Let’s see how you're doing first and then we can see when we can get you back to work; hopefully soon. How have things been since your surgery a week ago?”

PATIENT: “I've been doing okay. It’s been kind of painful. I was having a lot of pain in the largest wound. It was rather sore, so I took the oxycodone that you gave me and after I had difficulty pooping. I eventually took Extra Strength Tylenol. This is why I hate taking any medications.”

DOCTOR: “Yeah, yeah, that makes sense. Uh, tell me a little bit more about the pain at the surgical site.”

PATIENT: “Uh, so it's sore. Everything else seems OK, the one in the belly button hurts.”

DOCTOR: I’m sorry. Where exactly does it hurt?”

PATIENT: “This big wound by my belly button hurts a lot.”

DOCTOR: “That was just the instrument we used to operate. And is the pain there all the time? Does it go away at certain times during the day?”

PATIENT: “Look, I work in construction, and there is a deadline coming up to install this pool. If I don’t get it done, I won’t get paid.”

DOCTOR: “I understand and want to clear you for work, but I need to ensure you are doing well with the surgery. Is the belly button pain worse when you move at all?”

PATIENT: “Sometimes I feel like it's pulling. Other times, it is like pressure.”

DOCTOR: “When is it pulling? And have you noticed any redness or swelling around that area?”

PATIENT: “When I move around. There is no redness or swelling.”

DOCTOR: “OK. Well, that's good. And is the pain sharp or dull? Does it feel like it's, you know, you're being stabbed?”

PATIENT: “Well, you did make an incision to do the surgery, right?”

DOCTOR: “Yes, we did. Are you saying that it is sharp?”

PATIENT: “It is dull.”

DOCTOR: “Well, yes, I did make a small incision {PATIENT speaks simultaneously...} to insert a camera and surgical instruments. I closed the incision.”

PATIENT: {Upset tone} “You mean large incision...”

DOCTOR: “That being said, you can still feel pain. Have you had any fevers while you've been at home?”

PATIENT: “Uh, no.”

DOCTOR: “OK so no chills either?”

PATIENT: “No.”

DOCTOR: “Have you noticed anything coming out of the incision itself? Any fluid that's coming out.”

PATIENT: No, no fluid coming out. Maybe initially there was like a little bit of water from the belly button, I guess, but after that it was better, but more recently I have been having loose stools."

DOCTOR: "What do you mean by that?"

PATIENT: "Yeah, I've been having a lot of watery diarrhea."

DOCTOR: "Oh, OK, alright. And then just one last question about the incision and we could talk about the diarrhea. Have you noticed any bumps around there? Like maybe a fluid collection under the incision or anything?"

PATIENT: "Uh, no."

DOCTOR: OK, that's good. And you said you're having diarrhea. How long has that been happening?"

PATIENT: "Initially she told me to take stool softeners. It helped me go to the bathroom, but now it is watery and keeps going after I stopped the stool softener."

DOCTOR: "How many times are you pooping a day?"

PATIENT: "Uh, so initially pretty often probably like four to five times."

DOCTOR: "OK and it's still really watery?"

PATIENT: "Yeah, very watery."

DOCTOR: "OK. And that's not typical for you? Before the surgery, you were having pretty normal bowel movements, you'd say?"

PATIENT: "Yeah, yeah."

DOCTOR: "OK. How are you eating?"

PATIENT: "Well, actually I'm eating what I used to eat."

DOCTOR: "OK, what does that typically look like for you? Can you run me through a few meals during the day?"

PATIENT: "Ohh you know like probably like a morning sausage sandwich with eggs and some cheese. For lunch, I have like a burger with some more cheese on top and maybe french fries and a milkshake, you know? And then for dinner I try to eat a little lighter, you know... Some fried chicken and you know, potatoes, mashed potatoes and things like that."

DOCTOR: "Wow, those things sound really good. Well, I think uh, you know, we'll talk a little bit more about that.
It sounds like there are some changes we can make to your diet that might help you out with the diarrhea that you're having. But before we do that, let's go ahead and take a look at your incisions and at your belly and make sure that everything looks good there. OK."

PATIENT: "OK"

DOCTOR: "Let me take a look at your chart. It looks like your weight is stable. Your temperature is good. You don't have a fever. Your heart rate is in the normal range. It's only 76 and your blood pressure looks pretty good, 126/76, as well, and you've been taking your blood pressure medication, right?"

PATIENT: "Uh. Yep."

DOCTOR: “OK, great so let's see. Everything looks good there and we didn't get any labs for you. And I don't think that we need them right now, but let's go ahead and take a look at your belly. Your abdomen feels nice and soft and it doesn't seem like you're distended, and that's a good thing because you've been having diarrhea. So we want to make sure there aren't any problems there. OK, let's take a look at the incisions. They look good. I see this is the one that hurts, right? This biggest one here.”

PATIENT: “Ohh yeah, that hurts.”

DOCTOR: “Yeah, well, that makes sense. The incision by the belly button is the biggest incision. We went in there with small instruments and a camera to look at your gallbladder and then we're able to get it out of this incision.
This incision is the one that we take the gallbladder out of and it was filled with a lot of stones. So, let's take a look here. It doesn't look like there's any fluid that's collected under the incisions.”

DOCTOR: “It doesn't look like there's any pus or anything coming out of the belly button incision. It is a little bit red around there, but that's pretty normal for an incision still healing.”

PATIENT: “Are you trying to open the incision again...??”

DOCTOR: “I was just trying to ensure that there is no separation or breakdown. So yeah, it looks good. It doesn't look like it's infected at all to me.”

PATIENT: “That makes sense.”

DOCTOR: “I think it’s probably a good idea to come back in a week to see if the redness and your pain improve.”

PATIENT: “So I can go back to work?”

DOCTOR: “Let’s go back to the diarrhea you were having. That's actually a pretty common thing that we hear from our patients after they get their gallbladder out. Usually that's because you're having a little bit of a change in your body in the way that you digest things.”

DOCTOR: “So what we tell patients is that we'd like them to cut back on foods that have a lot of fats. It sounds like you have a lot of things like cheeses and French fries and fried foods. You might need to cut back on them, at least for the next few weeks.”

PATIENT: “I know.”

DOCTOR: “While your body makes adjustments do you think that's something that we can do?”

PATIENT: “I can't have cheese in my diet? I love swiss cheese, cheddar cheese, mozarrella, goat cheese, and my favorite, feta. My mom makes the best cheesecake. You didn’t tell me I must give this up before the surgery. Shouldn’t that be discussed as a risk to the procedure?

DOCTOR: “No, I get that. --
I love cheese, too, but just for a few weeks. And you should be able to uh, you know, start having cheese, again. You could introduce it back slowly, but for now, because you're having the diarrhea, it's probably a good idea to cut back on it.

{Interrupt}
PATIENT:
-- “What if I keep my diet but take the oxycodone that you gave me? That should balance things out.”

DOCTOR: “Let me get back to your question about the oxy. You said you don’t like taking medications when you don’t need to. Do you think you're able to manage the soreness around that incision without any oxy?”

PATIENT: “Yeah, as long as I can go back to work.”

DOCTOR: “In that case, I do not recommend the oxy anymore. Please just the Extra Strength Tylenol as needed.”

PATIENT: “So I can eat the other stuff. I just can't eat cheese and the French fries.”

DOCTOR: “That's right. And none of this will really hurt you if you're eating it. It's just, it sounds like it's pretty uncomfortable to be going to the bathroom several times a day and having diarrhea.”

PATIENT: “No, it’s fine. It is only once or twice a day now. But I will cut down the fatty food I am eating.”

DOCTOR : “So for the next couple of weeks, I would stick to more vegetables, things like that. And then after that slowly start to resume some healthy cheese. You know, it's good in general to try to cut back on those things, but you know, we're gonna eat what we're gonna eat.”

PATIENT: “Alright, sounds good.”

DOCTOR: “Well, that's good. And then it seems like overall you're doing okay. For your construction, do you do a lot of heavy lifting and things like that?”

PATIENT: Typically, I supervise, but you know, sometimes my guys need help working or like someone doesn't show up. So, I have to do some of the work too.”

DOCTOR: “So I think it's probably OK for you to go back to work now, since you're not really taking any pain medications or anything like that, but I would hold off on lifting anything for another couple of weeks. So typically we say 4 weeks out of surgery, you don't want to be doing any heavy lifting, anything more than a gallon of milk. So I would hold off on doing that if you're just around the construction site supervising and talking to people, that's fine.”

PATIENT: “OK, that's fair. Can you give me a note for that?”

DOCTOR: “Yeah. We can do that. And then, otherwise, your incision looks good. It's healing well, but we do want to keep an eye on it. If you start getting fevers, chills, notice some discharge, or if there is fluid collecting under it, then I want you to send me a message in the portal.”

PATIENT: “Ok will do.”

DOCTOR: “And then we already talked about the diarrhea and what we could do about that and hopefully that gets better once you change your diet.”

PATIENT: “I hope so too.”

DOCTOR: “Alright. Well, do you have any other questions for me?”

PATIENT: “Can I give the oxycodone to my aunt who is dying of cancer?”

DOCTOR: “Well, I am sure that your aunt has her own prescription. I would just drop the oxycodone at your local pharmacy so it can be disposed of safely. Do you have any other questions?”

PATIENT: “No, thank you.”

#### 3.5. Case 5

{Loud background noise, machines beeping, etc,.}

ED ATTENDING (Mat): “Okay guys what we have?”

NURSE-1 (Franny): “Bob, can you document? This is my patient.”

NURSE-2 (Bob): “Sure thing.”

EMS Provider: “I have a 53-year-old male who was involved in a MVC. He was unrestrained and his car hit a pole.  The guy is really lucky. He is mostly complaining of chest pain and trouble breathing. We placed in an IV and gave him some fluids. We slapped on a non-rebreather and he is oxygenating 100%. His blood pressure has been fine.”

NURSE-1: “What size IV did you place?”

EMS Provider: “Come on, Franny. You know me... It’s a trauma. Of course, I placed in an 18 gauge.”

NURSE-1: “Which arm?”

EMS Provider: “Left.”

ED ATTENDING: “What was his initial vital signs on scene?  What are the patient's last vital signs.”

EMS Provider: “Uh, let me look here.  So his heart rate was 120 beats per minute.  His blood pressure was 80/50, but like I said I gave him fluids and it is now 95/63.  His respiratory rate was about 20 respirations per minute.  His oxygen saturation was 80% on room air but like I said I gave him oxygen through non-rebreather and now it is about 92%.”

ED ATTENDING: “Let’s move him over to our stretcher. Charles, start your assessment.”

NURSE-2: “Sorry, can you give me those vitals again?”

EMS Provider: “Blood pressure was 96/53. I gave him some fluids and now, it is 80/50.  His respiratory rate was about 30 respirations per minute.  His oxygen saturation was 80%.

ED INTERN (Charles): “Sir, sir, what is your name?”

PATIENT (Gerry): “Gerry”

ED INTERN: “Can you tell me what happen?”

PATIENT: “Yeah… I.. I.. I.. can’t.”

EMS Provider: “The guy is just drunk. He smells of alcohol.”

PATIENT: “I really can’t breathe…”

ED ATTENDING: “I want a set of vitals”

ED INTERN: “Airway is clear... Breath sounds present but diminished on the left... I feel some crepitus on this chest... Good pulses bilaterally.”

NURSE-1: “Heart rate is 140 beats per minute. Blood pressure is 95/63. Respiratory rate is 40 breaths a minute. Oxygen saturation is 90% on 15 liters per minute with non-rebreather.”

PATIENT: “I... can’t... breathe...”

ED INTERN: “Patient has a small frontal hematoma... Pupils are 4mm bilaterally and reactive... There is no tenderness over the face...”

ED ATTENDING: “Charles, the FAST exam comes first.”

ED INTERN: “OK. Turning on the ultrasound.”

NURSE-1: “There is an 18 gauge IV placed in the right arm.”

PATIENT “I... can’t... breathe...” {Patient is agitated}

ED INTERN: Sir, please stop moving. The patient is possibly drunk and is agitated. Please get some Ativan.”

ED ATTENDING: “Wait, I believe that the patient has an impending tension pneumothorax.”

ED INTERN: “Can we get x-ray here?”

ED ATTENDING: “We need to decompress the chest, quickly. Franny page the surgeon. Put on sterile gloves and place the sterile drapes on the patient. Give me lidocaine 1% with epinephrine and give the patient fentanyl 50mcg IV. Charles, find the 4th intercostal space in the mid-axillary line and inject the lidocaine. You will need to inject in all directions and then into the pleural space.”

NURSE-2: “Doctor, did you want to proceed with the fentanyl? The patient’s blood pressure is 95/63.”

ED ATTENDING: “Good point. Let’s hold off.”

ED INTERN: “Ok.”

ED ATTENDING: “Clean the same area with betadine with outward concentric circles. Give me a scalpel. Take this scalpel and make a cut along the 5th rib. Make the cut big... like 3 centimeters.”

ED INTERN: “Ok.”

ED ATTENDING: “Take the Kelly clamp and use blunt dissection and inward pressure until you reach the pleura. Blunt dissection means to intermittently open the clamp to make the incision bigger.”

NURSE-1: “Triage is asking you to evaluate this 90-year-old lady on anticoagulation who fell and hit her head.”

ED INTERN: “I am at the --
pleural space.”

{Interrupt}
ED ATTENDING:
-- “Please tell the triage nurse ask another ED doc to evaluate that patient? I am in the middle of a procedure.”

ED ATTENDING: “Apply a lot of force but cautiously --
And you will feel a pop.”

{Interrupt}
ED INTERN:
-- “I am through. There is a gush of air.”

ED ATTENDING: “Open the Kelly one last time to make the incision bigger. Place your finger in the incision and remove the Kelly. Sweep your finger around to ensure there are no adhesions, and you can feel the lung.”

ED INTERN: “Ok.”

ED ATTENDING: “Take this 32Fr thoracostomy tube and place it in the incision. You are going to slide the tube posteriorly and to the apex. Stop when the fenestrations are in the chest. Franny, bring the drainage system and turn on the suction. It looks like there is blood as well. Charles, suture the tube in place.”

{SIM overhead: “CODE BAT”}
ED INTERN: “Ok.”

ED ATTENDING: “I need repeat vital signs”

NURSE-1: “Heart rate is 140 beats per minute. Blood pressure is 100/80. I think the surgeon is calling...
Respiratory rate is 20 breaths a minute. Oxygen saturation is 97% on 15 liters per minute with non-rebreather.”

ED ATTENDING: {on phone} “Hi, Randeep. I have this 53-year-old guy who was an unrestrained driver and sustained injuries after an MVA when he hit a pole. EMS reported initial hypotension but fluid responsive. Here, his shock index is greater than 1 and he was found to have an impending tension pneumothorax and we decompressed the chest. Post-thoracostomy tube, vitals are: Heart rate is 130 beats per minute. Blood pressure is 100/80. Respiratory rate is 20 breaths a minute. Oxygen saturation is 97% on 15 liters per minute with non-rebreather. My concern is that there is blood coming from the tube, which is measuring about 750mL so far. Please repeat the blood pressure.”

NURSE-2: “The chest x-ray was completed.”

ED ATTENDING: “His x-ray of the chest has multiple rib fractures on the left side. He has significant subcutaneous emphysema. The chest tube is in the correct place.”

PATIENT: “I feel really thirsty... Can I get water?”

NURSE-1: “Blood pressure is 70/40.”

ED ATTENDING: “The patient is in hemorrhagic shock. Activate massive transfusion protocol now. Randeep, I think this patient needs to go to the OR. I need you to come now. I will admit it to your service.”

ED INTERN: “I placed xeroform dressing on after I sutured the chest tube in place. I did the rest of the exam. FAST was negative. Abdomen is soft and non-tender. There were no other injuries. He is just weak overall but moving his arms and legs. We rolled him and there is no tenderness of his back.”

NURSE-1: “The first unit of whole blood is being transfused in the rapid infuser.”

{Trauma surgeon comes in}
TRAUMA SURGEON (Randeep): “It looks like the chest tube put out 1500mL of blood. We need to take him to the OR. Can we take him to CT before going up? What is blood pressure?”

NURSE-1: “Two units are in... Blood pressure is 90/40.”

TRAUMA SURGEON: "Let's keep giving blood products, take him to CT, and go to the OR."

ED ATTENDING: "Gerry, you suffered a tension pneumothorax from a car accident and required an emergent thoracostomy tube. You have blood in the chest, called a hemothorax, which is most likely from a blood vessel. Since it is continuing to bleed causing hemorrhagic shock, you required multiple blood transfusions. You may have other injuries from your car accident and you need a CT scan which the surgeon will review the results once it is obtained, but you need to go to the OR emergently with Dr. Singh, the surgeon who is at the bedside, to ligate the bleeding vessel."

TRAUMA SURGEON: "Hi, Gerry. I will take you to the OR, and you will be admitted to my surgical ICU."

#### 3.6. Case 6

DOCTOR: "Hi, Mr. Smith. I'm Doctor Tang, and I'm one of the inpatient hospitalists, and the ER Doctor told me that they'll be admitting you today. Is that OK with you?"

PATIENT: "Is that what he said? Oh man, Jack, is that true? Is that what he said when he left?"

DOCTOR: "Hi Jack. That's what they told me. Is that what they told you?"

SON (Jack): "Yeah, exactly. So, I am his son. From what I heard, he has some sort of an infection, and he needs some sort of antibiotic. I'm not too sure. The last time this happened, he just took a pill and got better. What's going on, Doc?"

DOCTOR: "OK. Well, it seems like I'll be admitting him to figure out exactly what is going on. I've read the charts so I can go over a couple of things together but they're basically admitting you for diabetic foot infection and we need to give you antibiotics. How I'd like this to go is that I need to do some charting and some paperwork. I need to ask you some standard questions. Then I'll put in all the appropriate orders, get you a meal, get you the antibiotics, put you back on all your home meds and then call the appropriate specialist to get you better. I will review the lab work with you and answer any questions you have at the end. Is that OK?"

PATIENT: "Fine, go ahead. I mean, it's already 1:00AM in the morning. This is like the third time I tell my story but whatever.

DOCTOR: "Alright, OK so. So, you're here for the foot infection. Can you tell me when it started? When was the last time that you were feeling well, and you were feeling fine?"

PATIENT: "What day is today? Saturday morning? So, I think sometime last week I would guess. I mean, I was doing fine doing well with no problems and taking my meds. Jack here was helping me out. He's been giving me the pills and giving me my insulin shots. You know, just doing good overall. This past Monday I felt something on my big toe. My cousin, Donna, had something similar, I think an ingrown toenail and she just cut it out. At first, I thought it was nothing, so I just cut my toenails that day."

DOCTOR: "Is that on the right side or the left side?"

PATIENT: "I was on my right big toe. So that was kind of it or so I thought. On Tuesday it was kind of getting worse. I couldn't really walk so I took off my shoe after gardening a bit and I told Jack to take a look. He said there was some sort of red thing under the big toe, and there might have even been pus then, so he got kind of worried. He knows I have diabetes. We went to the urgent care Tuesday, and they gave me some pills. Not too sure of the name. And you know, we thought it would get better."

DOCTOR: "So you went to urgent care and you saw a doctor there. What did they say? What was the problem?"

PATIENT: "I didn't really pay attention. Talk to Jack over there, he knows more."

DOCTOR: "Jack, what did they say? I'm sorry we have to repeat some stuff. I just need to make sure that we get everything correct. What did they tell you at the urgent care center?"

SON: "Sure, Doctor Tang. Let me back up a little bit and tell you what the story is. So this past week my dad was doing well. He was doing gardening but on Monday he mentioned something about his toe feeling a bit off, but he didn't really ask me to take a look at it or anything. On Tuesday he really started to bother him, and you know, when I took a look, the toe itself was kind of red. When I pressed on it, it hurt. It was swollen and there might have even been some pus. So, we actually went to the urgent care, and they gave him an antibiotic.
It started with the letter B. I want to say B. Help me out here doc."

DOCTOR: "Is it Bactrim?"

SON: "Yeah, yeah, that one. So, he took it. He took his antibiotics like they told him and on Thursday I took a look at it again and it wasn't getting any better --
and actually looked even worse."

{Interrupt - Annoyed}
PATIENT:
-- "It was getting actually getting better."

SON: "Dad....you can't see well without your glasses. Your vision is bad…It looked more red, more swollen and it was more tender. My dad really hasn't been walking since then because of the pain. I was bringing him breakfast, lunch and dinner every single day so we took him to the urgent care again Thursday and they kind of popped it with a needle and I think liquid started coming out. It was yellow and they changed the antibiotic to, I think, doxycycline. We gave it most of Friday but it looked worse and so we came in last night to the hospital. He has this diabetes, we know sometimes people get bad infections with diabetes, so we decided to get it checked out here in the hospital."

DOCTOR: "OK. I'm glad you made that decision and came to the ER. I think you know you're being admitted to the hospital, so we need to be a little bit more aggressive with therapies to get you better. Let me ask you, it sounds like you have diabetes. How long have you had diabetes? And do you know what your last hemoglobin A1C is?"

PATIENT: "I don't know, Jack, what do you think, 30 years? Probably since Jack was around five years old. When I was first diagnosed, they started with pills and now they have me putting these shots in every day. I keep having to check my fingers and look at my fingers they're all calloused now."

DOCTOR: "Who's your primary care doctor. Dr. Jones?"

PATIENT: "Yes. Dr. Jones."

DOCTOR: "Got it. Does Dr. Jones follow your lab work? What was your last hemoglobin A1C?"

PATIENT: “That’s the one you check every three months, right? The level is good.”

DOCTOR: “Yeah, that one. Do you remember the value?”

PATIENT: “Oh, it is 11. That’s good, right?”

DOCTOR: “11, That is not ideal. And what are your sugars at home usually?”

PATIENT: “I don't usually check to be honest with you. Jack here would know. Why don't you ask him?”

DOCTOR: “Jack. What is your dad's fingerstick glucose levels typically?”

PATIENT: “He usually never wants me to check. I’d say it’s in the 200-250’s range a good day. Sometimes, he won't let us check. Dr. Jones says he should take it three times a day. But you know what can III do?”

DOCTOR: “Alright, I understand. And do you take a long-acting insulin as well?”

PATIENT: “Long-acting. Is that the one with the L? I think Lispro.”

DOCTOR: “I think you mean Lantus.”

PATIENT: “That's it. That's it.”

DOCTOR: “All right. And have you guys been on any trips recently?”

PATIENT: “No.”

DOCTOR: “Any trauma to the legs?”

PATIENT: “To be honest with you, I love to garden. And sometimes I don’t wear sandals when I garden. I might have hit my foot, what, like two weeks ago? We were having a BBQ and gardening. Maybe something around then I’m not entirely sure.”

DOCTOR: “Got it. Do you have any history of neuropathy of your legs?”

PATIENT: “What's that?”

DOCTOR: “It is where you have altered sensation. For example, are your feet numb?”

PATIENT: “Always. I’ve had that for so many years, I take this one medication, gabapentin.”

DOCTOR: “Got it.”

PATIENT: “It doesn't help at all. I can still barely feel my toes.”

DOCTOR: “OK, alright. And do you see a podiatrist?”

PATIENT: “Yes.”

DOCTOR: “When was the last time you saw a podiatrist?”

PATIENT: “I’d say about 2-3 years ago.”

DOCTOR: “OK.”

PATIENT: “The podiatrist told me to follow up every year, but I haven't had time.”

DOCTOR: “Fair enough. Any bug bites, animal bites, anything like that?”

PATIENT: “Oh, nothing like that.”

DOCTOR: “So, let's get back to what brought you here. So, it looks like this started this past Monday, and since then, it has progressively gotten worse. You started on Bactrim on Tuesday and then on Thursday prescribed doxycycline, but despite this, your symptoms have gotten worse, and that's why you're here today. Does that sound about right?”

PATIENT: “Exactly.”

DOCTOR: “OK. And now I'm gonna do something called the review of systems. I'm gonna go from head to toe just to make sure that we didn't miss anything important. Just tell me if it's yes or no, OK. So, do you have any headache?”

PATIENT: “No.”

DOCTOR: “Chest pain? Difficulty breathing?”

PATIENT: “No shortness of breath. I have a cardiologist. I always get that pain in my chest. You know, they even did that stent thing on me like, what, five years ago, and I still have symptoms.”

DOCTOR: “Maybe we can go over it.”

PATIENT: “When I walk too much, I definitely get that chest pain still.”

DOCTOR: “OK. Let's finish the review systems first, and then we'll circle back to this chest pain. You said you didn’t have difficulty breathing. Any abdominal pain?”

PATIENT: “No.”

DOCTOR: “Diarrhea? Constipation?”

PATIENT: “To be honest with you Doc, since I started these meds on Tuesday, I think I've been having some of that. Yeah.”

DOCTOR: “Sometimes that happens with antibiotics. OK, so you have a little bit of diarrhea. Would you say you have bloody diarrhea, or is it watery?”

PATIENT: “Yeah, something like that. I’ve been going three times a day since I started the antibiotics. I’d say it's just watery.”

DOCTOR: “Got it so three times a day. OK, and you do have right toe pain, right?”

PATIENT: “Yes.”

DOCTOR: “OK. So, the three main things in the review systems are right toe pain, diarrhea, and chest pain. Does that sound correct?”

PATIENT: “Yeah, chest pain. When I walk. Doc, when I walk a lot.”

DOCTOR: “OK. Let's dig in a little deeper with the chest pain. So when did the chest pain start?”

PATIENT: “For the last 20 years. I noticed it getting worse and worse, and that's what made me wanna go to the cardiologist about 10 years ago and told me I might have cardiac issues. I didn't really follow up too much. Five years ago, I went to the cardiologist again, and the doc made me get a test. It was called a coronary scan and they thought I had some sort of blockage there, so I went under and they put in a stent."

DOCTOR: "Do you happen to know where they put the stent?"

PATIENT: "Where was it again Jack?"

SON: "It was the left anterior descending dad, the widowmaker they called it."

PATIENT: "Oh, that's the one. That's the one."

DOCTOR: "OK. Did they put in any other stents?"

PATIENT: "Oh, that's it."

DOCTOR: "What medication do you take?"

PATIENT: "I was taking two meds for a while for that, but now it's just a baby aspirin."

DOCTOR: "You don't take any blood pressure medications?"

PATIENT: "Oh yeah, that too. Lisinopril 10 milligrams every day."

DOCTOR: "Oh, you take lisinopril for blood pressure. Anything else? What about for cholesterol?"

PATIENT: "I have that too. Yeah, I've been trying to eat healthier and I'm trying to do more gardening and outdoor things. Doc I have so much going on. I have the toe thing, this chest pain, this neuropathy of my legs, I just don't like walking since this started because of the toe pain."

DOCTOR: "What do you take for cholesterol?"

PATIENT: "Oh yeah, the cholesterol. Jack, what was it? Let me take a look here. Hold on one second, its rosuvastatin 40mg every day."

DOCTOR: "Got it thanks."

PATIENT: "Isn't it in the computer already, Doc? Its 1:00 AM in the morning…."

DOCTOR: "I apologize. I want to make sure we have this right so we can treat you optimally while you are here in the hospital. So in the computer I have 40 milligrams orally at bedtime. Sounds like the dose you had mentioned."

PATIENT: "Yep."

DOCTOR: "OK. All right. That's what looks like what was prescribed and sent to a pharmacy three weeks ago. So, I'm figuring out what else you take. The other medication that I see is lispro for meals and that you take 25 units of Lantus at night. Is that correct?"

PATIENT: "Yeah, doc, that sounds about right."

DOCTOR: "OK. So going back, after the stent was placed, did you feel the pain was finally relieved or did it continue afterwards?"

PATIENT: "Tell you the truth, it's probably 80% better than five years ago right before the stent. But like I said, after walking more than 10 minutes, it definitely bothers me."

DOCTOR: "And to clarify you said you haven't gone back since then, right?"

PATIENT: “I haven't gone back to the cardiologist since the catheterization and stent placement.”

DOCTOR: “So since you've got your catheterization, you never went back.”

PATIENT: “Exactly. I just take this baby aspirin.”

DOCTOR: “OK. And when was the last heart test that you had? Like an echocardiogram?”

PATIENT: “Five years ago a few months after the stent was placed.”

DOCTOR: “Got it. Do you feel any chest pain right now?”

PATIENT: “Not usually but I do, talking to you right now.”

DOCTOR: “OK. I know it’s late, and hopefully, we can make you feel better. I forgot to ask you on review of systems, but have you felt fevers at all?”

PATIENT: “No.”

DOCTOR: “No fevers. Good. Now I'm gonna do a physical exam. I'm gonna speak my physical exam out because I'm gonna use my app here to help me write the note if that's OK with you.”

PATIENT: “Whatever you want, Doc.”

DOCTOR: “From the top, it looks like on your head I don't feel any fractures or anything like that. Your head is normocephalic and atraumatic.
Your pupils are equal and reactive.
Your tongue is midline.
Your oral mucosa is moist.
Your heart is regular rate and rhythm with no murmurs, rubs, or gallops.
Your lungs are clear to auscultation bilaterally.
Your abdomen is soft and non-tender in all four quadrants, with bowel sounds in all four quadrants.
You have mild right lower extremity edema of one plus with erythema on your right great toe, which radiates up to your midfoot. Your right toe is tender to palpation.
On your left, you have no edema, and you have no erythema nor swelling.
You have no focal neurological deficits.
Did I forget to ask you what your medical problems are? I think I did. What are your medical problems? I know that you have heart disease, and I know that you have diabetes and high cholesterol. Anything else that I’m missing?”

PATIENT: “Something about my kidneys. They say my kidneys aren't working too well. It's like 1.9 or something like that.”

DOCTOR: “They’re probably referencing your creatinine level. Anything else? It seems like you have something called CKD. Any other medical problems?”

PATIENT: “I don't think so.”

DOCTOR: “My chart says here that you have a blood problem.”

PATIENT: “Blood problem? Jack? Have you heard about this blood problem before?”

DOCTOR: “It’s called MGUS. Have you heard about it? Maybe your Primary Doctor is just observing at this time?”

SON: “About 10 years ago, my dad went to an oncologist/hematologist and said he had something in his blood.
But you know, we went every year, and they just kept checking it and have never said anything much about it. Should we be worried?”

DOCTOR: “No, no. You have something called monoclonal gammopathy of uncertain significance. Usually, there's nothing to do except to monitor it. So as long as you're following with somebody, you can continue to follow with them. Do you have any allergies?”

PATIENT: “I wanna say penicillin.”

DOCTOR It's important to know that you're allergic to penicillin so we can put you on the right medicine. And what is your reaction to penicillin?”

PATIENT: “I get itchy.”

DOCTOR: “OK. But there's no difficulty breathing, no tongue swelling, or anything like that, right?”

PATIENT: “No, I’d just power through it if I take it again.”

DOCTOR: “OK got it. As far as social history goes, I'm gonna have to ask some of these questions. Do you drink alcohol?”

PATIENT: “Not anymore.”

DOCTOR: “All right. When did you stop, and how much were you drinking?”

PATIENT: “Since that heart attack? I’d say five years ago. I was drinking maybe 3 beers a day.”

DOCTOR: “But no alcohol now, right?”

PATIENT: “No.”

DOCTOR: “And then what about smoking? Have you ever smoked?”

PATIENT: “No.”

DOCTOR: “Any illegal drug use?”

PATIENT: “Absolutely not, doc.”

DOCTOR: “Good. As far as surgical history, you had that one coronary stent. Any other surgeries in the past?”

PATIENT: “They took out my gallbladder like 9-10 years ago.”
“
DOCTOR: “So that's about 2014. Any family history, any illnesses with your mom, dad, brothers, or sisters?”

PATIENT: “My parents passed away from old age, and I'm an only child.”

DOCTOR: “Alright, so let's review things. So, I'm gonna show you the X-ray that was done. It shows some inflammation of the right toe. Xrays are usually very general studies for a possible localized infection that you may have.”

PATIENT: “You think you'll have to amputate the toe?”

DOCTOR: “Well, I don't think we're at that point yet, but we're gonna do a couple of tests and involve some specialists to see where we go from here.”

PATIENT: “Got it.”

DOCTOR: “There is some inflammation swelling in the right toe area which we can see on exam. Let me just bring up your lab work real quick.
Your vital signs are fine.

Your blood pressure's a little high. It's 159 / 79 with a heart rate of 79. Your temperature is 38.2 Celsius.
But your oxygen saturation's great and you're 99% on room air now.
I see here your sodium's fine at 139.
Your potassium is 4.9.
Your kidney function is elevated with a BUN level of 49, and your creatinine today is 1.9.
It seems to be consistent with what you had on prior visits. It seems your creatinine is typically around 1.3 to 1.5. So you do have something called CKD, but it's stable.
You do have an elevated white count of 15.30.
Our normal WBC count is around 10 and you do have a little bit of anemia, which you've had before.
Your hemoglobin here is 12.5.
You're usually around 12.4. The normal value for someone is 14.
So you've been anemic for a little bit, but it hasn't changed too much.
Other than that, we have two other markers of inflammation, something called the ESR. The ESR is elevated at 45 and your CRP is 1.5 which is also elevated.
So, you know all this points towards something called a diabetic foot infection. The ER doctors have already given you doses of two antibiotic medications called Zosyn and Vancomycin.
We want to see how deep the infection is and whether it's affected the bone because we want to rule out something called osteomyelitis. So, our first step in the diabetic foot treatment is going to be to continue the current antibiotics. We're going to get an MRI of your foot to rule out osteomyelitis. After we'll also get an arterial duplex of your legs on the right side to make sure that that you have good blood flow to the toe. Sometimes poor blood flow to the toe leads to poor wound healing. I'm also going to do a venous duplex ultrasound to rule out clots in your legs called dvt so you'll be going for a bunch of leg scans. From there we'll have our podiatrist and our infectious disease doctors come by. And out of curiosity, when the urgent care doctor popped the fluid, did they get something called a wound culture?”

PATIENT: “I think they did something with it. You're right, they popped it. They took a deep swab and they said that the results will be back in a couple of days and they they'd call me. So far no one has called me.”

DOCTOR: “When we finish, I'll get that number from you and have my people call the urgent care and see if we can get the results because that can help us figure out what the best antibiotics will be.”

PATIENT: “Right.”

DOCTOR: “So, as I said, we're going to be calling our infectious disease and our podiatrists. We may also ask for our vascular surgeon to come in if we find anything on some of those scans that need treating. The next problem on the list is your chest pain. It is kind of concerning that you've had continued chest pain for the past five years, and the fact that it's on exertion, it could indicate an active process. I only have one EKG here from the ER, and it doesn't look like you're having an ST elevation MI on it. On the EKG, you have what we call a first-degree heart block, and you have some PVCs, so I will get some bloodwork called a troponin level.
So we already drew your blood once. I'm going to be drawing it at least two more times while you're here to make sure that you're not having an active heart attack. We might get an echocardiogram, as you're long overdue for one, and have our cardiologist come see you. That's not an urgent thing, though, unless the troponins come up elevated.
The next problem on your list is that you're having this diarrhea. I suspect that it's something due to the antibiotic doxycycline OR Bactrim that you have been taking recently. Those medications can be associated with a superinfection called C diff. The next time you have poop, I will send the test to check for that. You know, you do have a bit of a fever, and you do have an elevated White Count, but your belly's not tender, but I want to rule it out.

The other problems seem to be more chronic medical problems. We're gonna continue treating you for your high blood pressure. We're gonna continue treating your cholesterol and check your lipid panel as well and continue your home medications. If we recheck your hemoglobin A1C and it is more than 11, I'm going to have you see one of the endocrinologists and a diabetic educator to see if we can get you back on track. Ideally, you'd be closer to 6.5, as that can really affect how fast you get better.
The next couple of questions I'm gonna ask are just routine questions we ask everybody. The first one is your activity level. You said that prior to having this infection, you were able to walk. Did you need a walker or cane or a wheelchair or anything like that or did you walk without an assistive device."

PATIENT: "I walk without any sort of support. It's just that after 10 minutes of walking that's when I really get the chest pain that I was telling you about."

DOCTOR: "Gotcha. OK. While you're here, I'm gonna make sure that the nurse is gonna help you out just in case, because you know, with the toe pain and the chest pain there should be someone there to help you out. We'll have our physical therapist come see how you are doing with walking."

PATIENT: "If I have to go to the restroom, can I go on my own or do I have to call for someone to help me up and walk?"

DOCTOR: "Press the button here and someone will come to help you up and to the restroom."

PATIENT: "OK got it."

DOCTOR: "The next question I had was regarding your diet. Do you have any difficulty eating foods at all?"

PATIENT: "No."

DOCTOR: "Great. I'm gonna put you on a cardiac diet at regular consistency. The final question we ask everybody when they come in to the hospital is your code status. If your heart stopped working and your lungs stopped working, would you want chest compressions? electric shocks? Would you want to be on a breathing machine? Is this something that you've thought about before?"

PATIENT: "Yeah, I talked to Jack about this. Definitely do all that you can to keep me alive."

DOCTOR: "OK. So that's called full code and I'll make sure to make a note of that in the chart. In case of emergencies, do you have a health care proxy?"

PATIENT: "Jack?"

DOCTOR: "Healthcare proxy's a legal form, and that's where you are assigning someone to take care of your medical decisions in cases where you can't make decisions for yourself. Right now, you're fully alert and oriented so you're not going to have to worry about that. But I do need to ask you if in case of emergencies, do you have a health care agent or someone that you want us to call if you don't have one, then just tell us who you wanna call in case of emergencies."

PATIENT: "Just call Jack."

DOCTOR: "Jack, what's your number?"

PATIENT: "(516) 123-4567"

DOCTOR: "Got it thanks. Any questions?"

PATIENT: “It's two in the morning on a Saturday Doc. Can you just turn off the light and let me sleep?”

DOCTOR: “All right well then, I know it's late. I'm gonna be here until 7:00 AM. I'll get all those orders in and let you both get some rest. Thank you both. You've been very helpful, and I appreciate you.”

PATIENT: “Bye.”
